# *“The early years are like a foundation for the future”* Perspectives, facilitators and challenges of Anganwadi Workers’ in supporting early child development interventions in Hyderabad, India: Qualitative findings from A Scalable Programme Incorporating Early Child Development Interventions (ASPIRE)

**DOI:** 10.1101/2023.01.11.23284414

**Authors:** Gitanjali Lall, Reetabrata Roy, Kunduru Sharath Chandrika, Gauri Divan

## Abstract

ICDS, a large public health programme, addresses the needs of children with ‘Anganwadi Workers (AWW)’ as frontline agents of delivery. This paper describes formative work done with AWWs, as part of ASPIRE to assess their understanding of Early Child Development (ECD) and acceptability of a novel ECD intervention. Framework analysis of their responses from FGDs led to identification of three themes:1) time use 2) understanding of ECD and 3) delivering messages using videos. The findings suggest that AWWs tight schedules, often leave them feeling overburdened with work. They are aware of factors that can aid as well as hinder child growth and development, but their understanding of play is limited to games played by older children. They expressed acceptability in using a video intervention, specifying features that would increase relevance for families. These have implications for ECD intervention development, training needs of AWWs as well as policymakers in the field.

**What We Already Know:**

- Frontline workers are one of the most effective agents for delivery of universal ECD interventions in the community, given their relationships with families
- Anganwadi Workers (AWW), are government frontline workers in India who work to improve the nutritional and health status of children, improve the capacity of the mothers to take care of their children’s health and nutritional needs and to support pre-school learning opportunities
- AWWs are overburdened with work and lack the time as well as proper facilities for intervention delivery

**What This Article Adds:**

- Integration of novel ECD interventions delivered by frontline workers need to take into account their existing work schedules and associated challenges
- Training on ECD interventions will need to broaden AWWs understanding of the critical foundational experiences which responsive caregiving and early child stimulation can provide
- The factors that are promoters in effective implementation of community-based ECD interventions

## Introduction

Early childhood development (ECD) has received significant global attention in the last decade^1^, with research across low- and middle-income countries (LMIC) showing that interventions delivered in the first three years of life are effective in improving ECD outcomes^2^. Despite this, millions of children under five years residing in LMICs are at risk of not reaching their development potential because of poverty, poor health, nutrition, and suboptimal care^3^.

India has responded to needs of young children through different approaches^4^, largest being the Integrated Child Development Services (ICDS) Scheme^5^. With children up to six years, pregnant and lactating women as its beneficiaries, the strength of ICDS is its ability to reach the remotest areas of the country^6^. The scheme is implemented through platforms called Anganwadi Centres (AWC) managed by Anganwadi Workers (AWW), mandated to support ICDS activities and associated administrative work^7^. An initiative implemented since 2018 in ICDS is Poshan Abhiyaan or National Nutrition Mission which strives to reduce rates of stunting, under-nutrition, anaemia and low birth weight by linking and monitoring different nutrition schemes for children^8^.

Any programme involving AWWs has to understand their time utilization, especially since they often deliver multiple programmes^9^. Research conducted on AWW time use and challenges of their work, using observations, interviews, or both^9–13^ has found that they are able to spend only 54% of their time on core responsibilities of pre-school education and home visits with substantial burden of record maintenance^10,11^. A third of the AWWs feel weighed down because of their involvement in national health programmes and election duties, in addition to routine responsibilities^9^. AWWs also report dissatisfaction with low honorarium, delay in receiving funds, inadequate infrastructure, scarcity of play materials, lack of regular refresher trainings and spending a significant amount of time ensuring that the AWC is clean and has drinking water^10–13^. Key challenges within ICDS include inadequate emphasis on behaviour change activities, infrequent home visits, lack of respect accorded to AWWs and insufficient focus on children in the zero-three years age^6^. As a result, ICDS in many parts of the country is being administered more as a “welfare support than (a) developmental activity”^4(p.232)^.

Studies in India have found implementation of integrated nutrition and early learning interventions through ICDS feasible^14^. A Scalable Programme IncorpoRating ECD Interventions (ASPIRE), developed by Sangath in collaboration with UNICEF and Department of Women Development and Child Welfare (DWDCW) Telangana intends to develop, implement, and evaluate a video based intervention integrating nutrition and responsive caregiving in the first 1000 days of life in the south Indian state of Telangana. The intervention will be embedded into ICDS and delivered by AWWs who are already trained on delivering messages via videos using the Common Application Software (CAS), which enables Real Time Monitoring (ICT-RTM) of service delivery under the Poshan Abhiyaan ^15^.

This study describes formative research conducted to inform the development and plan implementation of ASPIRE. It aims to understand (1) time-use of AWWs (2) perspective of AWWs on ECD (3) experiences of AWWs with video usage on CAS and (4) AWWs attitudes to delivering a novel ECD intervention.

## Methods

Focus Group Discussions (FGD) were conducted with AWWs from Khairtabad, Alwal and Maheshwaram ICDS projects in November and December 2019. Six FGDs, having five-six AWWs in each group were conducted with 31 AWWs. None refused consent to participate. Discussions were moderated by a Research Assistant with Masters in Public Health, well versed in reading, writing and speaking the local language (Telugu)[CK] and assisted by a Research Associate with Masters in Developmental Psychology, having working knowledge of Telugu [GL]. FGDs were conducted either at project offices or AWCs as per availability.

The interview guide and Participant Information Sheet (PIS) were developed in English and translated to Telugu. An initial pilot FGD conducted with AWWs allowed for refinement of the guide. The semi-structured FGD Guide addressed AWW time use including daily and monthly activities, knowledge of child growth and development, understanding of play, opinions on video interventions, challenges and facilitators to work.

Prior to the FGD, a presentation on ASPIRE was shown, following which contents of the PIS were discussed and written informed consent was obtained for all participants. After conducting the first three FGDs, findings were discussed with project principal investigators [GD, RR] and the guide was refined.

CK developed expanded notes upon completion of each FGD. Data was analysed using framework analysis^16,17^, which aimed to identify similarities and differences in the data set for extracting themes^16^ often referred to as framework categories^17^. CK and GL familiarized themselves with the expanded notes, identifying themes based on study objectives. Microsoft excel was used, with rows representing themes and columns representing responses; a final column summarised responses from all FGDs. Categories and subcategories are detailed in the next section. This work was approved by Sangath’s Institutional Review Board on 15^th^ October 2019.

## Results

### Participant demographics

Table 1 shows characteristics of the participating AWWs; as can be seen the mean age of AWWs was 41 years with an average experience of 16 years. Majority (48.4%) had an undergraduate degree as their highest level of education. The number of beneficiaries registered at each AWC varied greatly, with mean number of registered pregnant women being 14.7(5-25), lactating mothers 12.7(6-25) and preschool children 46.3 (22-75).

**Table 1:** Participant Characteristics (N=31)

| Characteristic | Mean (SD), Range |
| --- | --- |
| Number of years of experience | 16.1 (5.6), (8-29) |
| Age | 41.1 (6.4), (31-56) |
| Highest level of Education | Percentages |
| 10 Standard | 19.4 |
| 12 Standard | 12.9 |
| Graduate | 48.4 |
| Postgraduate | 19.4 |
| Number of registered beneficiaries at AWC | Mean (SD), Range |
| Pregnant Women | 14.7 (6.7), (5-25) |
| Lactating Mothers | 12.7 (5.3), (6-25) |
| Preschool Children | 46.3(17.9), (22-75) |

### Findings of Framework Analysis

Six framework categories, broadly divided into three major themes:1) AWW time use, including AWWs opinion of their rewards and challenges, 2) AWWs understanding of ECD and 3) AWWs views on delivering messages using videos, were identified. Key findings are described below

#### Anganwadi Worker Time Use

A key area of exploration was to understand what activities AWWs conducted and time allocated to each of these.

##### Daily and Monthly Activities

AWWs reported investing between six and nine hours a day on Anganwadi work, including pre-school education, provision of nutritional supplementation, home visits and documentation. AWWs reported adapting ICDS prescribed work schedules to allow for accommodating personal needs.

The first half of each day was focused on pre-school teaching. A prescriptive daily pre-school activity schedule designed by DWDCW was followed which included pre-school lessons, specific activity times for lunch, nap, snack and self-study sessions. AWWs reported seeing a drop in preschool attendance due to mandatory requirement of a national identification card which some children don’t have. Moreover, many parents preferred to get their children admitted to local private schools instead of AWCs, because of lower age of admission at private schools and their proximity to family homes. Additionally, given that children were between the ages of three and five, AWWs did not feel like they were able to manage a class of more than 20-25.

During preschool hours, eligible beneficiaries (pregnant women and lactating mothers) for supplementary nutrition (‘*Aarogyalaxmi*’) are mandated to eat their food at the AWC. During this time, AWWs stepped away from preschool activities to address issues on antenatal care. Six out of 31 AWWs reported being unable to spend quality time with the women due to engagement in preschool activities. AWWs also described a drop in quality of food being supplied since transitioning from freshly cooked food to that being supplied by charitable trusts.

*Ayamma* (Anganwadi helper) *cooking at the AWC is good because people from the trust used to cook food in the morning and it becomes cold till it reaches here. And the taste wasn’t good. Ayamma washes the vegetables two-three times and cooks in front of the beneficiaries*

-FGD 6

After completing preschool activities, AWWs conducted home visits for pregnant women and children under two years. They reported being able to conduct up to four visits each day using *‘Intintiki Anganwadi’* (House to House Anganwadi), a home visiting guide containing age-specific content on nutrition, stimulation and play.

Their usual work day ended between 5:30 and 6pm.

Besides daily activities, AWWs described organising additional weekly and monthly events including the Nutrition Health Day (NHD) or Village Health Sanitation and Nutrition Day (VHSND-1), Immunization Day targeting children or VHSND-II, *Swaraksha* Day, Early Childhood Care Education (ECCE) Day and Anganwadi Level Monitoring and Supporting Committee (ALMSC) Meeting. AWWs were also required to conduct two Community Based Events every month on Saturday afternoons based on themes provided by DWDCW. These included children’s birthdays, hand-wash days and *‘Annaprasanna’* (first complementary feed) day, usually combined with other monthly events.

These activities are detailed in Table 2

**Table 2:** Summary of Anganwadi Workers schedule of activities

| Daily activities |  |
| --- | --- |
| **General** | AWWs invest between 6 and 9 hours on designated Anganwadi activities every work day. |
| **Pre-school activities** | AWWs follow the daily pre-school activity timetable given by the Directorate, Women and Child Development, Telangana State<br>Daily timetable activities are often overridden by |
|  | monthly activities (see later). |
| **Nutrition**<br>**Supplementation** | • Eligible beneficiaries for supplementary nutrition include pregnant women, lactating mothers and pre-school children.<br>• 20-30% of the registered women pick up their nutrition supplementation from the AWC.<br>• Rice, Dal, and one boiled egg is provided at lunch for pregnant, lactating women and pre-school children to eat at the AWC.<br>• Eggs are given at home, three times a month for children between 7 months and 3 years of age.<br>• Pregnant and lactating women visit the AWC for a hot meal |
| **Documentation** | • 14 registers are maintained for daily activities and 6 registers for monthly events.<br>• 4-8 registers are completed daily.<br>• 4 registers are written once a month.<br>• Each register takes 15-20 minutes for completion, except the home visit register, completed during the home visits which takes about 30 minutes.<br>• During the research period, AWWs were entering date in both the paper and digital format due to the ongoing CAS pilot. |
| **Home Visits** | • AWWs conduct their home visits with pregnant, lactating women and mothers with children below 2 years once they finish with preschool.<br>• AWWs go for 1-4 home visits daily accounting for 15-75 home visits per month. It takes between 10 minutes and half an hour for each home visit. AWW's use the 'Intintiki Anganwadi' book to counsel beneficiaries and their family.<br>• For AWWs having CAS, the schedule for home visits shows up on the phone. |
| **Monthly Activities** | • Nutrition Health Day (NHD) /Village Health Sanitation and Nutrition Day (VHSND-1): 1<sup>st</sup> & 5<sup>th</sup> of every month; Anthropometry of all children and pregnant women. Distribution of 'balamrutham'; a fortified powder that contains iron, calcium, vitamins <sup>30</sup> for children between seven months and three years.<br>• Anganwadi Level Monitoring and Supporting Committee (ALMSC) Meeting: Conducted for a day between 1<sup>st</sup> and 10<sup>th</sup> of the calendar month at AWC; Discussions on stock at the AWC, food quality, pre-school activities and decisions on distribution of food at home to specific beneficiaries.<br>• Immunization Day (VHSND-2) held on a Saturday. In some AWCs, it is conducted twice a month |
|  | (additionally on Wednesday) if there is a large group of beneficiaries; Auxillary Nurse Midwives (ANM) give vaccinations to the children and pregnant women at the AWC.<br>• Swaraksha Day: 3rd Saturday during lunch break; AWWs visit schools in their catchment area to inform adolescent girls about personal hygiene, Helpline Numbers, Education, and Child Marriages. Occassionally this may be held in the AWC<br>• Early Childhood Care Education (ECCE) Day: 4th Saturday; feedback on each child's progress is given to mothers<br>• Community Based Events (CBE)*:(such as *Annaprasana*(first food), Children's Birthdays, Handwash Day): 2 events in a month based on themes given by DWDCW<br>• Distribution of eggs: thrice in a month |

##### Positives of AWW Work

Majority AWWs enjoyed preschool tasks, especially interacting and supporting teaching through play. One of them described a *‘sense of happiness’* in seeing children’s faces at the AWC. They took a sense of pride in their student’s achievements especially when they received reports of good performance in primary schools.

‘*Every year I send two-three Anganwadi children to primary school, their headmasters say those children from Anganwadi are gems. My children are always first in class*.’

FGD 3

They also felt respected when beneficiaries followed their advice.

##### Challenges of AWW Work

Majority AWWs reported unpredictable interferences in their work routines; these include duties assigned by other departments (e.g. distributing sarees for the local festival, election duty). The excessive workload, especially owing to duplication of efforts on completing registers on paper as well as digitally, often resulted in AWWs carrying work home. There were problems with infrastructure of AWCs, with many having insufficient space and lacking basic water and sanitation facilities, resulting in many AWWs spending their own resources towards maintenance of the AWC, for which they were not reimbursed. Over-scheduling of meetings every month, resulted in low beneficiary attendance. These systemic challenges meant that many AWWs felt dissatisfied with their salary, in terms of what they receive not being commensurate with their efforts.

Traditional beliefs that restrict movement of mothers who have recently given birth, outside homes interfered with their ability to receive food and supplementation from the AWC. There were also beliefs around the timing of introduction of complementary feeding, which often led to a delay in the same. This was aggravated by distance of homes from the AWC and fear of infections.

‘… *people from ‘Srikakulam’ (a district in Andhra Pradesh State) do not start complementary feeding till the age of nine months. They do not listen when we AWWs, ask mothers to feed the child from six months. They follow instructions given by their grandparents*.*’*

FGD 4

Since home visits were often delayed when parents were late picking-up their children, personal safety of AWWs became a key roadblock in conducting home visits during evening hours. AWWs also described feeling underappreciated, especially when parents accused them of not working, when they were out of the AWC to attend ICDS organised mandated events like trainings and meetings.

#### Understanding of Early Child Development

An understanding of early child development and growth amongst AWWs was sought, since monitoring this is an important mandated activity. Categories and subcategories which emerged are detailed below.

##### Conceptualization and Importance of Early Child Development

Growth was understood as weight and height for age. However, in two FGDS, a more expansive definition of growth as encompassing, ‘mind’ or ‘brain’ development emerged. A combination of nutritional factors (e.g. feeding children on time, exclusive breastfeeding until six months, followed by complementary feeding), environmental factors (e.g. neat and clean home), family characteristics (e.g. joint family) and good care during pregnancy supported growth. Factors associated with poverty viz., insufficient space, inadequate nutrition, and lack of playgrounds impeded healthy growth.

Development was described as age-appropriate achievement of milestones, which were supported by parents’ role (e.g. talking to child in the womb, parental knowledge about child development), home environment (e.g. a peaceful home where parents do not fight) and family characteristics (e.g. involvement of grandparents). Barriers to development included detrimental role of technology used as a distraction for children.

AWWs were able to recognize the importance of growth and development during early years, describing it as a ‘*basement of the house’* and ‘*investment for future generations’*.

*‘The early years are like a foundation/basement for the future* …*If we nurture them properly for the first two years…it will have a lifelong impact…*.*’*

FGD 2

##### Concept and Importance of Play

AWWs recognized the role of play in supporting growth, development and formation of friendships, and described games played at home and pre-school. These included outdoor games mostly for older children, fine motor activities, pretend play with a kitchen set, imitating the mother and teacher; and other activities such as making puzzles, building blocks and even playing on phone. Some examples of local games are described in Table 3. Responses to the age at which children start playing ranged from soon after birth to three years.

**Table 3:** Summary of findings (Understanding of ECD)

| **UNDERSTANDING OF ECD** |  |
| --- | --- |
| **Growth** |  |
| Facilitators | • Nutrition of the pregnant women and children including exclusive breastfeeding and timely introduction of complementary feeding |
|  | • Hygienic surroundings<br>• Child able to play and exercise<br>• Harmonious household<br>• Child growing up around grandparents<br>• Government and Community participation to ensure that individual recommendations to families are also implemented at the community level |
| Barriers | • Lack of good nutrition specifically, vitamin-d deficiency<br>• myths and beliefs in relation to food such as delayed introduction of complementary feeding<br>• Insufficient space for child to explore and play<br>• Financial difficulties in the family<br>• Mothers having inadequate knowledge of child development<br>• Lack of shared responsibility of childcare<br>• Family conflicts<br>• Genetics |
| **Development** |  |
| Facilitators | • Good care received by mother during pregnancy<br>• Talking to the child and enabling exploration by taking them outdoors<br>• Disciplining the child<br>• Allowing children to play games |
|  | • Adequate finances<br>• Peaceful home environment<br>• Clean and green surroundings<br>• Child growing up around grandparents |
| Barriers | • Mother's disturbed mental health during pregnancy<br>• Family conflict and violence<br>• Giving phones to children<br>• Parents unable to spend quality time with children<br>• Small families allowing lesser opportunity for interaction<br>• Physical environment: Small rooms allowing no space to play<br>• Myths and traditional beliefs which oppose current ICDS guidelines |
| **Games played by children locally** |  |
| Indoor games | • 'Poosalu Guchadam' (putting the pearls in a thread)<br>• 'Aksharala Chadarangam' (Alphabet game played on a board similar to chess),<br>• 'Ginjaluaata' (some seeds are put inside the circle and ask the children to blow. The children get more points if they blow more seeds out of the circle),<br>• chusthe cheptham chepthe thestham' (children |
|  | are encouraged to remove one object from the bag without seeing it and describe the object in their own words),<br>• ‘chintaginjal upper chatam’ (seeds are arranged in the form of flowers)<br>• Making clay or paper idols for local festival |
| Outdoor games | • ‘nichenaaata’ (Jumping on boxes drawn on the ground on with two legs),<br>• ‘Gili GiliAata’ (all the children hold their legs with their hands from the back and say the words *gili gili* and jump),<br>• ‘thadulodooradam’ (two children hold the rope on two ends and one child is asked to enter the other side without touching the head to the rope),<br>• ‘mundarakivenkakaki’ (Jumping forward and backwards)<br>• ‘buttalobanthi’ (throwing the ball in one basket). |

AWWs understanding of ECD is summarized in Table 3.

### Delivering Messages Using Videos

20 AWWs in two of three ICDS projects were using CAS to show video messages to beneficiaries and felt that this was effective in reducing their workload since it precluded the need to memorise messages; videos also have detailed and easy explanations of the topic being discussed and support standardised delivery. An AWW compared the effect of videos on people to local dramas, since it generated excitement in the viewer. Availability of very few videos however, had made the experience repetitive. Language of videos (Telugu) was also a barrier since it was not addressing the needs of a sizeable proportion of resident non-telugu speakers.

## Discussion

This paper explores time-use of AWWs, their perspective on ECD and experiences with video use in the South-Indian state of Telangana. The study was conducted as a formative, collaborative step in the development of a novel video based ECD intervention.

In line with previous research^9–11^, we found that AWWs seemed hard-pressed for time to complete their core activities, as a result of excessive workload arising from a lot of time spent on record keeping and non-ICDS related work. Some other challenges reported in the FGDs including inadequate or poor quality infrastructure, low honorarium and community perception that AWWs aren’t doing their work, have also been reported previously^10,18–20^. A novel finding, was that CAS, designed to ease this workload at least in the current piloting phase, seemed to be an additional burden. However, CAS is no longer being used and a new system is being introduced which addresses these drawbacks (Khyati Tiwari, MSc, e-mail communication, February 2022).

Among all of AWWs responsibilities, pre-school education was at the forefront: this is evident as the AWWs are referred to as ‘teacher’ in Telangana. Besides this, their responsibilities are focussed around nutrition, with minimal mention of play, or stimulation for zero-three year olds, despite this being a part of the state curriculum. This is consistent with findings from previous studies which have found AWWs possessing good knowledge on provision of nutritional supplementation^19–22^ and its facilitation in the community^18^, but demonstrate limited practice of early stimulation and learning activities for children, despite it being part of their service mandate^18^. The reduced attention to play and stimulation may be because government performance indicators such as the National Family Health Survey primarily measure nutritional status of children^23^, which is highly dependent on implementation of schemes by AWWs.

Previous research on knowledge of AWWs in ECD is limited and studies assessing their understanding of growth and its monitoring have yielded mixed results^24 22,25,26^. In this study, we found that AWWs understood growth largely in terms of height and weight, which was a reflection of their training on growth monitoring^7^. Development was understood essentially in relation to acquisition of milestones, which can be attributed to monitoring done using the Mother-Child Protection (MCP) card.

As in other studies, we noted that a number of AWWs interviewed, described ‘play’ as structured activities that happen with older children (See Table 3)^27^. Our novel findings are that there was awareness of parenting style and family environments which have potential of derailing optimal child development, reflecting AWWs recognition of key adverse childhood circumstances.

A strength of our study is the method of data collection used. Information from respondents was collected using focus groups, which enables collection of a full range of perspectives in a short duration, through a “group effect” in which participants’ responses augment one another^28(p.132)^. Ensuring confidentiality and using semi-structured interviews allowed AWWs to share their thoughts, especially in relation to health system challenges, freely. A key weakness of this study is the small number of participants from a limited geography which may not be representative of the larger population of AWWs.

We found that all AWWs were open to a novel ECD intervention with standardised video content. This may be attributed to the existing familiarity with the CAS, which despite its mixed perceptions amongst beneficiaries, has shown acceptability amongst AWWs and has demonstrated an improvement in quality of service delivery^29^.

An interesting observation made by AWWs was language diversity in the local population, given migration. This language difference between AWWs and caregivers, has caused communication difficulties impacting essential service delivery^18^; this needs to be a key consideration in design of any intervention.

## Conclusion

This study was undertaken as a formative step to inform development of a novel scalable video intervention for supporting optimal development and growth in the first 1000 days. Understanding time-use and ideas of development and growth of AWWs, helped us understand that the intervention once developed, will have to take into consideration challenges faced by AWWs with their current workload in order to be acceptable to them, while also acknowledging that the training will need to broaden AWWs understanding on the critical foundational experiences which responsive caregiving and early child stimulation can provide. Experience of AWWs and families with existing digital intervention, suggests that a variety of videos will need to be developed to be used as an assistive device rather than a replacement for messaging provided by AWWs.

In addition to direct implications for ASPIRE, the study provides insights into the experiences of frontline government workers in the DWDCW, Telangana which can be used to inform policy makers as well as researchers and interventionists in public health and ECD.

## Supporting information

Acknowledgements, Conflict of interest, funding

## Data Availability

The final instruments and all data are freely available with written permission from the authors

