## Supplementary material for "*“The early years are like a foundation for the future”* Perspectives, facilitators and challenges of Anganwadi Workers’ in supporting early child development interventions in Hyderabad, India: Qualitative findings from A Scalable Programme Incorporating Early Child Development Interventions (ASPIRE": Acknowledgements, Conflict of interest, funding

The authors of this study would like to express our deepest gratitude to our study participants for taking time out to discuss their views for the purpose of this study. We would also like to thank the Department of Women Development and Child Welfare, Government of Telangana for permitting us to interview the Anganwadi Workers and to UNICEF Office of Andhra Pradesh, Telangana and Karnataka for helping organise these permissions.

### Declaration of Conflicting Interests

The author(s) declare(s) that there is no conflict of interest.

The final instruments and all data is freely available with written permission from the authors.

### Funding

**The author(s) disclosed receipt of the following financial support for the research, authorship, and/or publication of this article:** This work was supported by the UNICEF Office for Andhra Pradesh, Telangana & Karnataka (PD No: PCA/2019/010, renewed subsequently as IND/PCA2019489/PD20221865 and IND/PCA2019489/PD20211299)
